# AI-VOICE: A Method to Measure and Incorporate Patient Utilities Into AI-Informed Healthcare Workflows

**DOI:** 10.1101/2024.09.19.24313990

**Authors:** Keith E. Morse, Michael C. Higgins, Yichun Qian, Alison Callahan, Nigam H. Shah

## Abstract

**Background:** Patients are important participants in their medical care, yet artificial intelligence (AI) models are used to guide care with minimal patient input. This limitation is made partially worse due to a paucity of rigorous methods to measure and incorporate patient values of the tradeoffs inherent in AI applications.

This paper presents AI-VOICE (Values-Oriented Implementation and Context Evaluation), a novel method to collect patient values, or utilities, of the downstream consequences stemming from an AI model’s use to guide care. The results are then used to select the model’s risk threshold, offering a mechanism by which an algorithm can concretely reflect patient values.

**Methods:** The entity being evaluated by AI-VOICE is an *AI-informed workflow*, which is composed of the patient’s health state, an action triggered by the AI model, and the benefits and harms accrued as a consequence of that action. The utilities of these workflows are measured through a survey-based, standard gamble experiment. These utilities define a patient-specific ratio of the cost of an inaccurate prediction versus the benefits of an accurate one. This ratio is mapped to the receiver-operator-characteristic curve to identify the risk threshold that reflects the patient’s values.

The survey instrument is made freely available to researchers through a web-based application.

**Results:** A demonstration of AI-VOICE is provided using a hypothetical sepsis prediction algorithm.

**Conclusion:** AI-VOICE offers an accessible, quantitative method to incorporate patient values into AI-informed healthcare workflows.

## Introduction

A core tenant of medicine is that patients should be empowered to be contributors to their own care. As artificial intelligence (AI) models become adopted into clinical practice, patients’ views should be considered when deciding how and if AI tools are used. Patient participation in the development and deployment of AI in healthcare is supported by the National Academy of Medicine^1^ and patient advocacy organizations.^2,3^

Well-meaning health systems attempting to honor this tenant are limited by a paucity of established methods to collect and incorporate patient values of the tradeoffs (i.e. benefits of accurate predictions versus the costs of inaccurate predictions) inherent in AI applications. While the medical decision-making literature has recognized the importance of such patient values since the 1970s, its adoption in evaluating AI applications is limited.^4^ Current approaches primarily rely on qualitative methods, such as focus groups or surveys, that evaluate patient perspectives on general considerations of AI in healthcare ^5,6^ or AI applied to specific disease domaines.^7–10^ Others advocate for the inclusion of patient representatives in the entire AI development lifecycle, yet details are lacking on specific contributions expected from the patient.^11,12^ Some quantitative methods exist that incorporate preferences into risk-based decision support, but often the preferences are collected from clinicians, not patients.^13^

Qualitative data collection methods are limited by cost infeasibility and poor actionability. Few health systems have the budget or methodological expertise to engage patient groups for each of the dozens of potential AI applications in their health system. Furthermore, once completed, it’s often unclear how such qualitative feedback can be concretely reflected in the implementation of the AI model. The end result is that patient values are rarely and opaquely included in AI-guided workflows.

This paper introduces AI-VOICE (Values-Oriented Implementation and Context Evaluation), a novel method to collect patient values in AI-guided workflows in healthcare. This method includes a survey that employs a standard gamble^14^ framework to quantitatively measure perceived value, or utilities, of hypothetical outcomes resulting from an AI-triggered action. The method can be applied to a diverse array of clinical or operational AI applications and is straightforward enough to be used by health systems without specialized expertise as part of routine model evaluation. The subjects completing the survey do not need specialized knowledge, thus is amenable to patient participation. Furthermore, the output of the survey is directly actionable by suggesting an optimum risk threshold at which follow up action should be taken (or halted). Thus this method offers an actionable path to infuse patient values into AI-guided care.

## Method

### Inputs

AI-VOICE is intended for use by hospitals or health systems that are implementing AI tools into their local care delivery processes. Here, the entity being evaluated is an *AI-informed workflow*, which is a combination of three elements: 1) the model and the risk estimate it provides, 2) the workflow and intervention triggered by the AI model, and 3) the near-term benefits and harms likely to accrue as a consequence of that intervention.

Use of AI-VOICE requires that the AI-informed workflow has already been specified, namely that a target clinical workflow has been identified, the AI model selected, and a policy developed that delineates what specific actions will be taken as a result of the output of the algorithm. The algorithm’s receiver-operator-characteristic (ROC) curve on appropriate validation data should also be available.

The research team should include a local subject matter expert on target workflow. For clinical workflows, that will likely be a physician with expertise in diagnosis and treating the target disease process. For non-clinical workflows, it will be an operational or organizational leader.

### Outcome Endpoints

The AI-informed workflow is described by a set of four outcome endpoints (OEs). OEs are text descriptions of the possible scenarios that could result in the AI-informed workflow and are extensions of a traditional 2×2 confusion matrix.

Whereas each quadrant of a 2×2 confusion matrix represents a ground truth and predicted class, each OE describes a ground truth state, an action triggered by the algorithm’s prediction, and a brief description of the near-term benefits and harms that accrue as a result of that action. Describing these benefits and harms are done by the subject matter expert and should be an attempt to describe the most likely expected clinical progression. Figure 1 shows the components of OEs.

**Figure 1:**
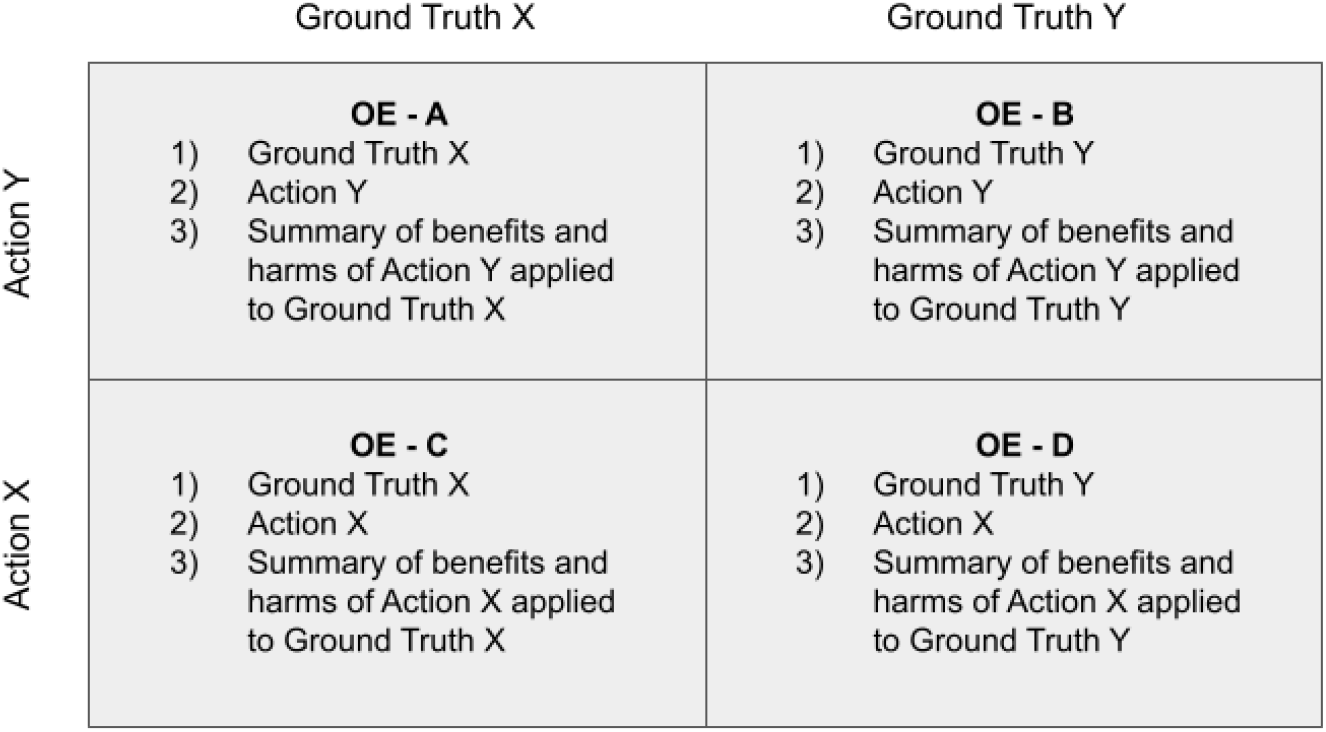
Components of outcome endpoints (OEs) for Actions X and Y, applied to Ground Truths X and Y

Note that OEs require substantial simplification of the complexities of medical care. The permutations of patient histories, disease severities, treatment efficacy and unintended complications means that no description could comprehensively include all potential outcomes of a disease course. The intention is not to be exhaustive, but descriptive of a single, representative disease course.

### Standard Gamble Experiment

AI-VOICE uses a standard gamble to estimate the patient utilities associated with each OE. Utility is a measure of health related quality of life derived from preferences that patients attach to their overall health status. Traditional methods to measure utility include standard gamble, time trade-off and willingness to pay^15^.

The first step to set up the standard gamble experiment is to rank the OEs in order of most to least desirable (i.e., highest to lowest utility). This is typically straightforward in clinical settings given that presence of disease or unnecessary treatment are less desirable than the absence of disease or necessary treatment. Note this method assumes that, in the presence of disease, treatment is preferable to no treatment and in the absence of disease, no treatment is preferable to treatment. Once ranked, the OEs are annotated with the highest utility as OE_1, down to the OE with the lowest utility as OE_4.

OE_1 is assigned a utility of 1 and OE_4 a utility of 0. The standard gamble experiment is then used to measure the utilities of OE_2 and OE_3 on the defined scale of 0 to 1.

Figure 2 lays out the basic structure of the standard gamble experiment. A participant is provided a choice between two options: Option A is a certain outcome of OE_2, and Option B is an uncertain outcome between OE_1 and OE_4. The probabilities of the uncertain outcomes is defined by *d* for OE_1 and 1-*d* for OE_4. The value of *d* is varied until the participant is indifferent between Option A and

**Figure 2:**
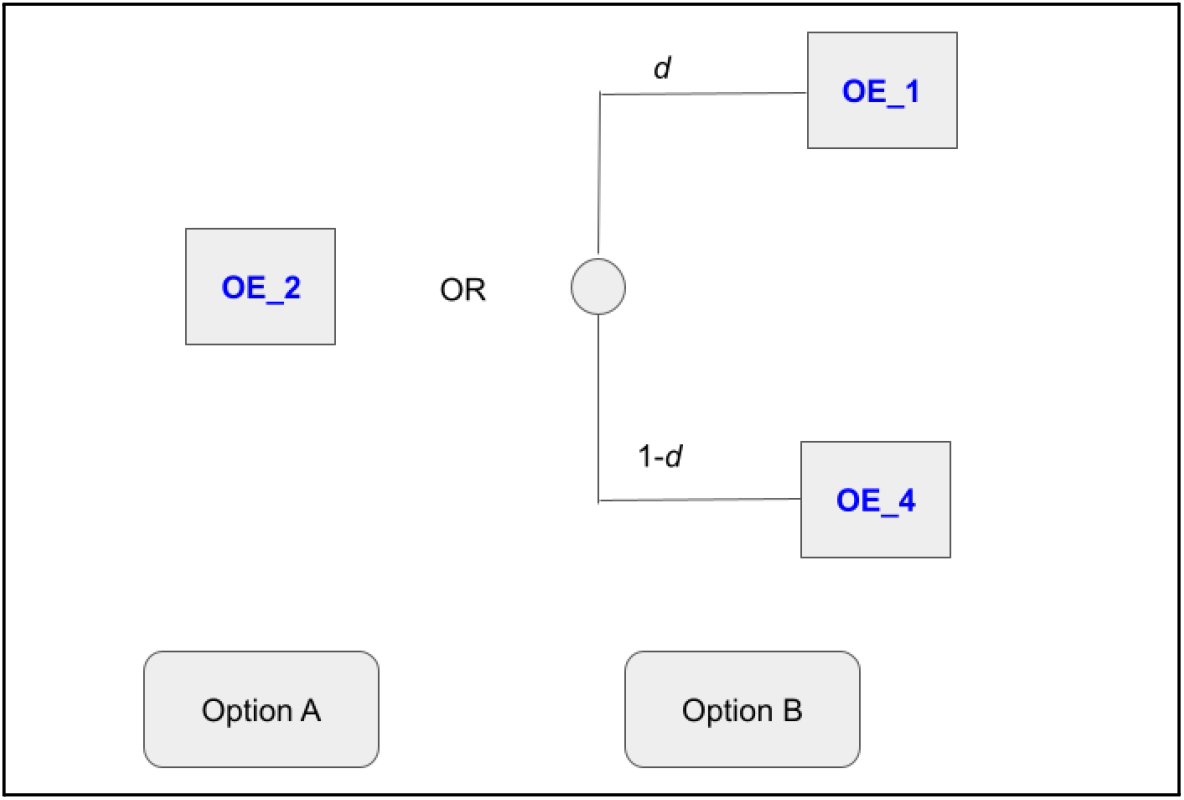
Standard gamble experiment to estimate the utility of OE_2

**Figure 3:**
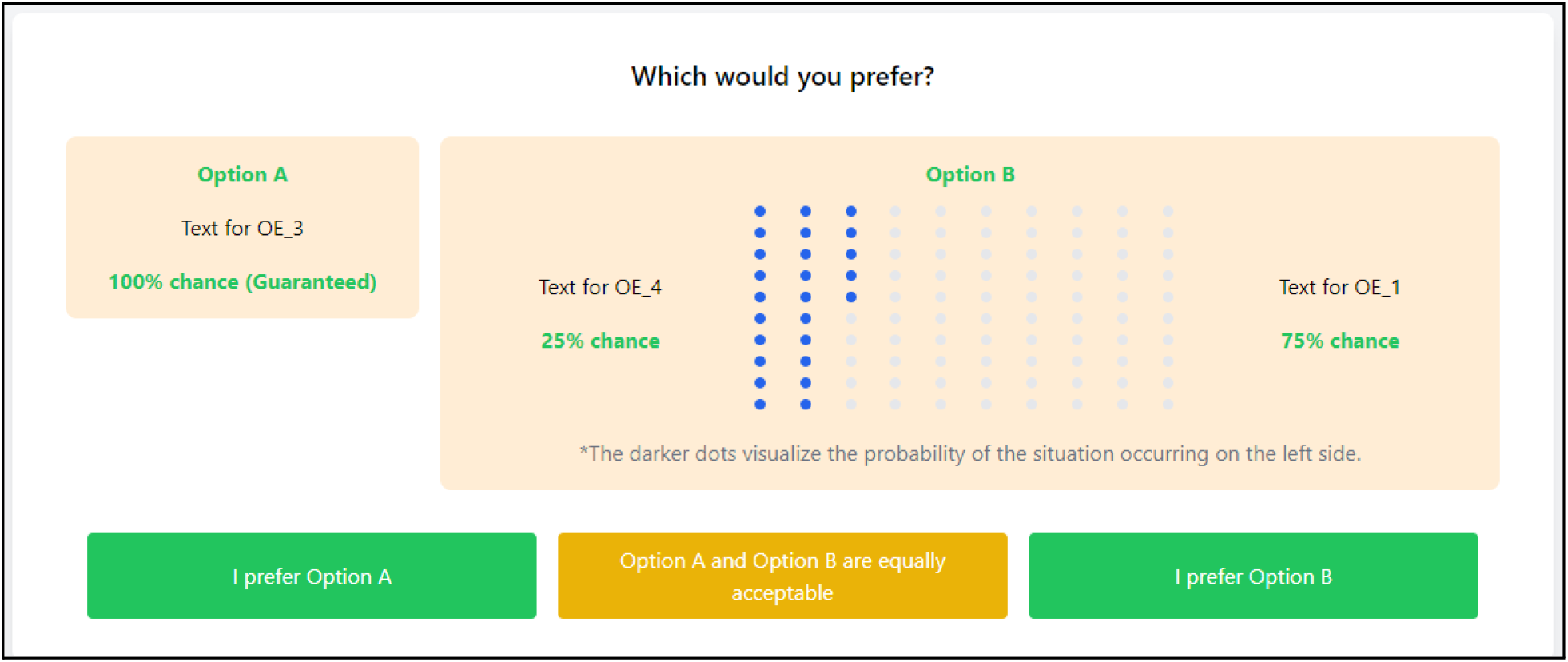
Screenshot from AI-VOICE survey instrument

**Figure 4:**
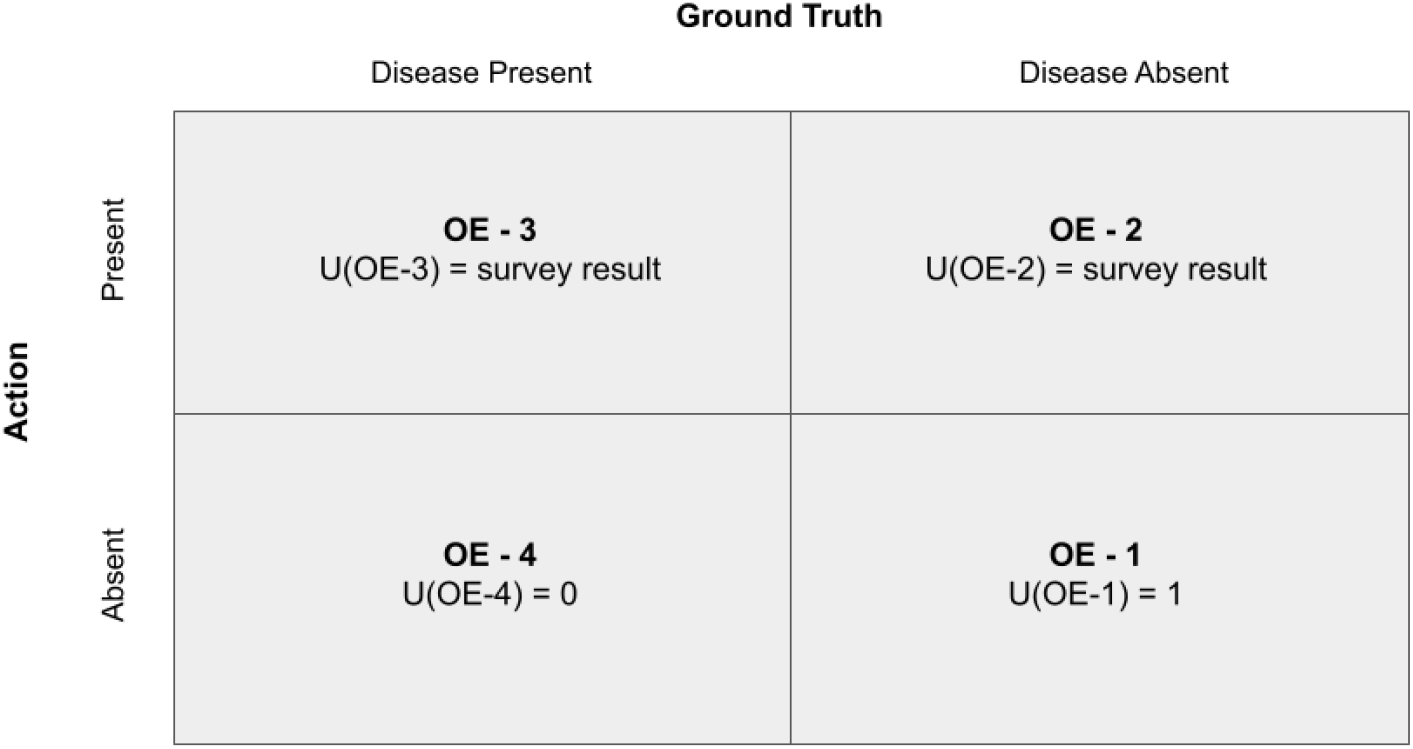
Utilities associated with each OE, after completion of surveys

**Figure 5:**
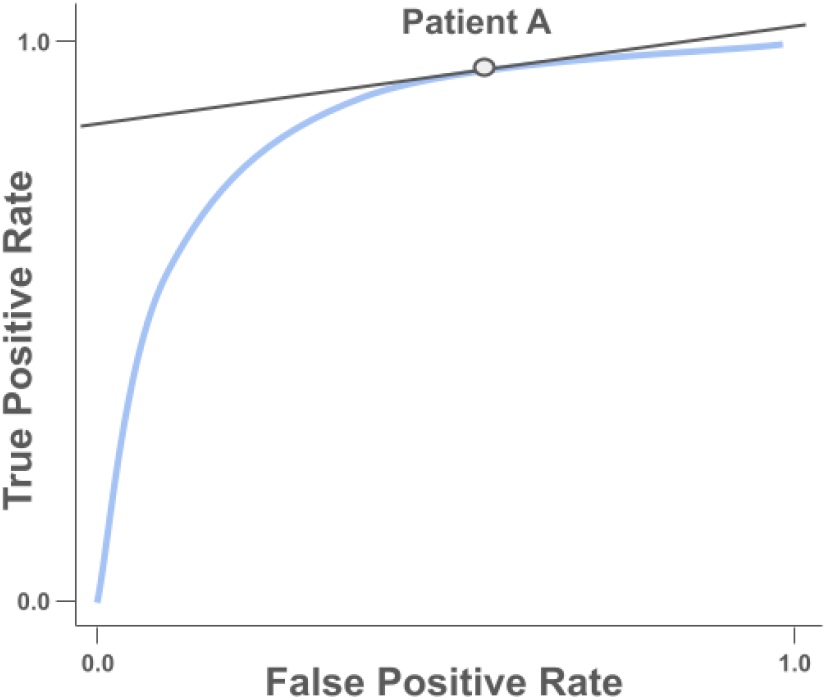
Patient As isopreference line used to identify optimal risk threshold on ROC curve

**Figure 6:**
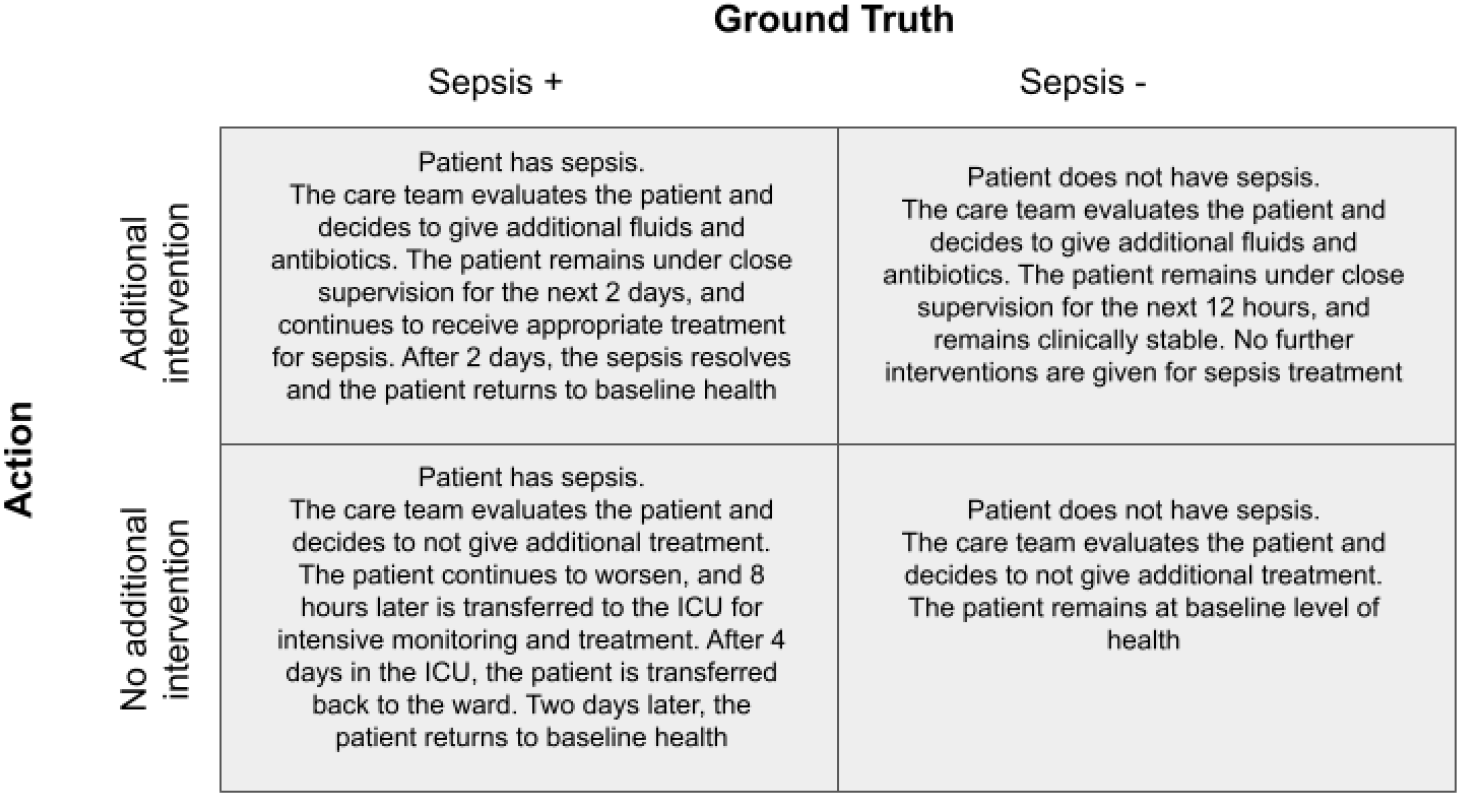
Outcome endpoints for hypothetical sepsis prediction algorithm

**Figure 7:**
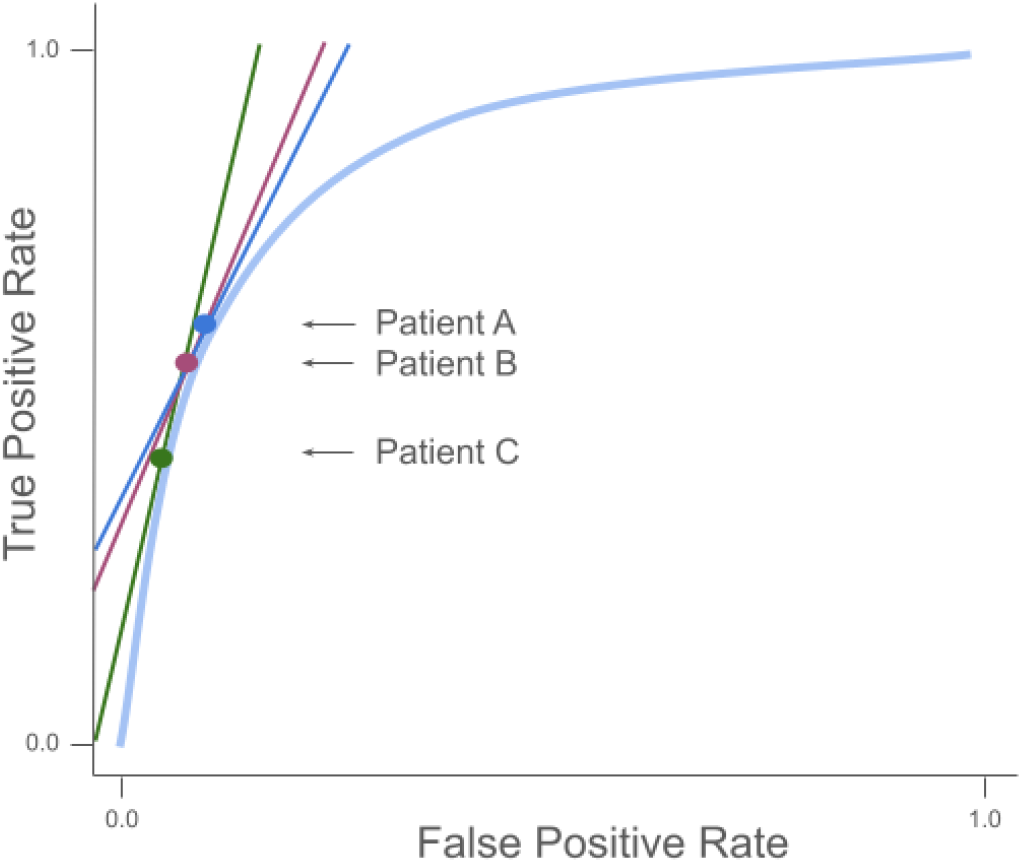
Sample isopreference lines plotted on ROC curve

Option B. At that point, the utility of OE_2(*U(OE_2)*) is estimated by:

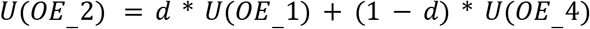

The above process is repeated to estimate the utility of OE_3, except where Option A is OE_3.

A web-based survey instrument to enable the development and distribution of the above standard gamble experiment to study participants is available at https://ai-voice.stanford.edu. This tool includes a researcher module that provides instructions for constructing OEs and ranking them. An access code is then available to be shared with study participants, who can complete the survey. A screenshot of a study participant’s view is below.

### Survey Results to Risk Threshold

The output of the survey are utility estimates for OE_2 and OE_3. Those, combined with the utility values of 0 and 1 assigned to OE_4 and OE_1, respectively, represent utility values for all OEs.

The next step is to translate the utility values to a risk threshold. Assume the optimal risk threshold is one that maximizes utility for the patient. The ROC curve can be seen as a function that maps the false positive rate (*f*) onto the true-positive rate (*t*(*f*)). The optimal false positive rate is the point on the ROC curve tangent to the slope defined by the equation below.^16^ See Appendix for detailed derivation.

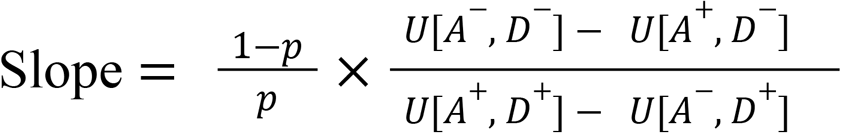

where

*p* = Pretest probability of disease

U[*A*^-^, *D*^-^] = Utility of action negative, disease negative scenario

U[*A*^+^, *D*^-^] = Utility of action positive, disease negative scenario

U[*A*^+^, *D*^+^] = Utility of action positive, disease positive scenario

U[*A*^-^, *D*^+^] = Utility of action negative, disease positive scenario

Recognizing that the *U*[*A, D*] terms equal the *U*[*OE*] terms, and replacing them in the equation gives:

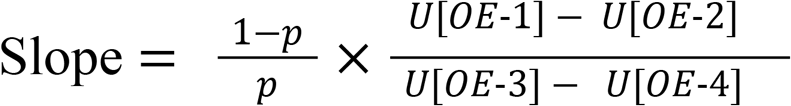

A value for *p* can be estimated from the model training data or known prevalence from local data. The *U*[*OE*] terms are input from the survey results. The resulting line, known as the *isopreference line*, identifies a point tangent to the ROC curve that is the optimal risk threshold given the patient’s values.

## Results

A demonstration of this method is presented using a hypothetical AI model and survey results.

The AI model is a sepsis prediction algorithm. For patients admitted to the inpatient ward, the algorithm evaluates vital signs, lab results and problem list every 15 minutes. For the patients deemed at high risk to develop sepsis in the next 2-6 hours, an alert is displayed in the electronic health record. Providers are expected to huddle with the bedside nurse to discuss the need for intervention, including the administration of fluids and antibiotics. The OEs for this algorithm are displayed in the below 2×2 table.

Illustrative results from the AI-VOICE survey from three patients are included in Table 1. A pretest probability of disease of 6% is used^17^ to calculate the isopreference lines.

**Table 1:** Sample results for three patients. Values in columns U[OE-2] and U[OE-3] obtained from the survey. Isopreference lines calculated using the provided equation.

| Survey Respondent | U[OE-2] | U[OE-3] | Isopreference Line |
| --- | --- | --- | --- |
| Patient A | 0.9 | 0.7 | **2.2** |
| Patient B | 0.9 | 0.6 | **2.6** |
| Patient C | 0.8 | 0.7 | **4.5** |

The isopreference lines are plotted below on an idealized ROC curve.

## Discussion

AI-VOICE is a method to quantitatively measure patient utilities of AI-guided workflows and translate those utilities into a risk threshold, thus offering a process to meaningfully incorporate patient values into AI-guide care.

The key patient information is represented in the above term 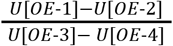. Here the numerator represents the net benefit that accrues to the patient *in the absence of disease*, as seen in the benefit of appropriately not being treated minus the cost of inappropriate treatment. Similarly, the denominator represents the net benefit that accrues to the patient *in the presence of disease*, as seen as the benefit of appropriate treatment minus the cost of inappropriate non-treatment. Taken together, this term can be viewed as an individualized trade-off between the benefit of appropriate treatment versus the cost of unnecessary treatment. Phrased another way, this term shows how many times a patient would be willing to be treated unnecessarily in order to receive needed treatment once.

The survey results can suggest instances in which an AI model is not needed at all. For isopreference lines near the extremes of zero or infinity, the added value of the model becomes negligible, and a policy of ‘treat all’ or ‘treat none’ is instead adopted. For example, an isopreference line near zero suggests that the cost of inappropriate treatment is exceedingly small relative to the benefit of needed treatment. In this case, a policy of ‘treat all’ serves the same purpose as an AI-algorithm with a low enough risk threshold that essentially everyone crosses it. Consider the example of getting an echocardiogram(ECG) to evaluate for the presence of a heart attack. The ECG is a quick, cheap, non-invasive procedure that imposes minimal cost on the patient. The benefits of identifying and treating a heart attack are substantial. Thus an AI model that identifies which patients should receive an ECG when there is concern for a heart attack will have an isopreference line close to zero, and can be scrapped in favor of giving ECGs to all patients with concern for heart attacks.

Each completed survey produces an isopreference line, and when multiple participants are surveyed, it is likely that multiple, different risk thresholds are identified. The process by which these competing risk thresholds are reconciled is beyond the scope of this work. Possible approaches include selecting the average risk threshold or choosing the most conservative value from the range of thresholds identified.

Further consideration is required if different risk thresholds are identified for different healthcare stakeholders. For example, if both patients and physicians are surveyed and the results suggest different optimum risk thresholds between the groups, selecting what risk threshold to use requires careful consideration. AI ethicists identify such ‘value collisions’ as prime sources of ethical problems, thus health systems should develop policies that acknowledge and address the ethical uncertainties in such AI applications.^18^

This method contains certain limitations. The first is that although patients do not require specialized expertise to participate in the survey, some degree of familiarity with medical terminology and care delivery is required to interpret the OE scenarios. Even if the scenarios adhere to guidelines that medical text be written at a 7th grade reading level,^19^ patients may have limited, or differing, understandings of the clinical experiences described. Further limitations stem from known weaknesses of the standard gamble methodology, including the high cognitive demands of considering complex probabilities used in the assessment, assumptions about equal risk tolerances among participants and the oversimplification of health decisions down to only two outcomes.^20,21^ Finally, extrapolating the values of a small number of survey participants to an entire patient population requires careful consideration of representative sampling methodologies.

## Conclusion

AI-VOICE offers an accessible, quantitative method to incorporate patient values into AI-informed healthcare workflows. Health systems adopting this method could improve patient engagement in their care.

## Data Availability

All data produced in the present work are contained in the manuscript.

## Bibliography

1. Adams L. Artificial intelligence in health, health care, and biomedical science: An AI code of conduct principles and commitments discussion draft. NAM Perspect. 2024;4. doi:10.31478/202404a

2. Light Collective - Patient AI Rights Initiative. Light Collective. Published March 22, 2024. Accessed August 21, 2024. https://lightcollective.org/patient-ai-rights/

3. Moore B. NHC Statement on Artificial Intelligence and Health Care: Promise and Pitfalls. National Health Council. Published February 8, 2024. Accessed August 21, 2024. https://nationalhealthcouncil.org/letters-comments/nhc-statement-on-artificial-intelligence-and-health-care-promise-and-pitfalls/

4. McNeil BJ, Weichselbaum R, Pauker SG. Fallacy of the five-year survival in lung cancer. N Engl J Med. 1978;299(25):1397–1401.

5. Musbahi O, Syed L, Le Feuvre P, Cobb J, Jones G. Public patient views of artificial intelligence in healthcare: A nominal group technique study. Digit Health. 2021;7:20552076211063682.

6. Richardson JP, Smith C, Curtis S, et al. Patient apprehensions about the use of artificial intelligence in healthcare. NPJ Digit Med. 2021;4(1):140.

7. Yap A, Wilkinson B, Chen E, et al. Patients perceptions of artificial intelligence in diabetic eye screening. Asia Pac J Ophthalmol (Phila). 2022;11(3):287–293.

8. Armero W, Gray KJ, Fields KG, Cole NM, Bates DW, Kovacheva VP. A survey of pregnant patients’ perspectives on the implementation of artificial intelligence in clinical care. J Am Med Inform Assoc. 2022;30(1):46–53.

9. van der Zander QEW, van der Ende-van Loon MCM, Janssen JMM, et al. Artificial intelligence in (gastrointestinal) healthcare: patients’ and physicians’ perspectives. Sci Rep. 2022;12(1):16779.

10. Nelson CA, Pérez-Chada LM, Creadore A, et al. Patient perspectives on the use of artificial intelligence for skin cancer screening: A qualitative study: A qualitative study. JAMA Dermatol. 2020;156(5):501–512.

11. Adus S, Macklin J, Pinto A. Exploring patient perspectives on how they can and should be engaged in the development of artificial intelligence (AI) applications in health care. BMC Health Serv Res. 2023;23(1):1163.

12. Banerjee S, Alsop P, Jones L, Cardinal RN. Patient and public involvement to build trust in artificial intelligence: A framework, tools, and case studies. Patterns (N Y). 2022;3(6):100506.

13. Vickers AJ, van Calster B, Steyerberg EW. A simple, step-by-step guide to interpreting decision curve analysis. Diagn Progn Res. 2019;3:18.

14. Thoma A, Bayoumi AM. Standard Gamble. doi:10.1016/B978-0-12-803678-5.00457-4

15. Bakker C, van der Linden S. Health related utility measurement: an introduction. J Rheumatol. 1995;22(6):1197–1199.

16. McNeil BJ, Keller E, Adelstein SJ. Primer on certain elements of medical decision making. N Engl J Med. 1975;293(5):211–215.

17. Rhee C, Dantes R, Epstein L, et al. Incidence and trends of sepsis in US hospitals using clinical vs claims data, 2009-2014. JAMA. 2017;318(13):1241–1249.

18. Char D, Abràmoff M, Feudtner C. A Framework to Evaluate Ethical Considerations with ML-HCA Applications-Valuable, Even Necessary, but Never Comprehensive. Am J Bioeth. 2020;20(11):W6–W10.

19. Mishra V, Dexter JP. Comparison of readability of official public health information about COVID-19 on websites of international agencies and the governments of 15 countries. JAMA Netw Open. 2020;3(8):e2018033.

20. Blinman P, King M, Norman R, Viney R, Stockler MR. Preferences for cancer treatments: an overview of methods and applications in oncology. Ann Oncol. 2012;23(5):1104–1110.

21. Beresniak A, Russell AS, Haraoui B, Bessette L, Bombardier C, Duru G. Advantages and limitations of utility assessment methods in rheumatoid arthritis. J Rheumatol. 2007;34(11):2193–2200.

