## Appendix for "AI-VOICE: A Method to Measure and Incorporate Patient Utilities Into AI-Informed Healthcare Workflows"

$U[T^{+}, D^{+}] =$ Utility of test positive and disease present

$U[T^{+}, D^{-}] =$ Utility of test positive and disease absent

$U[T^{-}, D^{+}] =$ Utility of test negative and disease present

$U[T^{-}, D^{-}] =$ Utility of test negative and disease absent

For when below conditions are met:

$U[T^{+}, D^{+}] > U[T^{-}, D^{+}]$

$U[T^{-}, D^{-}] > U[T^{+}, D^{-}]$

$p =$ probability of disease

$u(f) =$ expected utility for choosing cutoff point $f$ and $t(f)$

$$u(f) = (U[T^{+}, D^{+}] \times (t(f)) \times p) + (U[T^{-}, D^{+}] \times (1 - t(f) \times p) + (U[T^{+}, D^{-}] \times f \times (1 - p)) + (U[T^{-}, D^{-}] \times (1 - f) \times (1 - p))$$

$K_1$, $K_2$ and $K_3$ are constants relative to $f$ and $t(f)$:

$$K_1 = p \times (U[T^{+}, D^{+}] - U[T^{-}, D^{+}])$$

$$K_2 = (1 - p) \times (U[T^{+}, D^{-}] - U[T^{-}, D^{-}])$$

$$K_3 = (p \times U[T^{-}, D^{+}]) + ((1 - p) \times U[T^{-}, D^{-}])$$

The goal is to find the value of $f$ that maximizes

$$u(f) = K_1 t(f) + K_2 f + K_3$$

$x^*$ is a local maximum for $F(x)$ implies $\frac{dF(x)}{dx}\Big|_{x=x^*} = 0$

Therefore,

$$\frac{du(f)}{df} = K_1 \frac{dt(f)}{df} + K_2 = 0 \Rightarrow \frac{dt(f)}{df} = -\frac{K_2}{K_1}$$

Substituting values for $K_1$ and $K_2$ yields,

$$\frac{dt(f)}{df} = \frac{1-p}{p} \times \frac{U[T^-, D^-] - U[T^+, D^-]}{U[T^+, D^+] - U[T^-, D^+]}$$
